# Pre-Operative Kidney Biomarkers and Risks for Death, Cardiovascular and Chronic Kidney Disease Events: The TRIBE-AKI Study

**DOI:** 10.1101/2021.12.13.21266784

**Authors:** George Vasquez-Rios, Dennis G. Moledina, Yaqi Jia, Eric McArthur, Sherry G. Mansour, Heather Thiessen-Philbrook, Michael G. Shlipak, Jay L. Koyner, Amit X. Garg, Chirag R. Parikh, Steven G. Coca, TRIBE-AKI Consortium

## Abstract

**Background:** Soluble tumor necrosis factor receptor (sTNFR)1, sTNFR2, and plasma kidney injury molecule-1 (KIM-1) are associated with kidney events in patients with and without diabetes. However, their associations with clinical outcomes when obtained pre-operatively have not been explored.

**Methods:** The TRIBE-AKI cohort study is a prospective, multicenter, cohort study of high-risk adults undergoing cardiac surgery. We assessed the associations between pre-operative concentrations of plasma sTNFR1, sTNFR2, and KIM-1 and post-operative long-term outcomes including mortality, cardiovascular events, and chronic kidney disease (CKD) incidence or progression, ascertained after discharge.

**Results:** Among 1378 participants included in the analysis with a median follow-up period was 6.7 (IQR 4.0-7.9), 434 (31%) patients died, 256 (19%) experienced cardiovascular events and out of 837 with available long-term kidney function data, 30% developed CKD. After adjustment for clinical covariates, each log increase in biomarker concentration was independently associated with mortality with 95%CI adjusted hazard ratios (aHRs) of 3.0 (2.3-4.0), 2.3 (1.8-2.9) and 2.0 (CI 1.6-2.4) for sTNFR1, sTNFR2 and KIM-1, respectively. For cardiovascular events, the 95%CI aHRs were 2.1 (1.5 – 3.1), 1.9 (1.4 – 2.6) and 1.6 (1.2 – 2.1) for sTNFR1, sTNFR2 and KIM-1, respectively. For CKD events, the aHRs were 2.2 (1.5 – 3.1) for sTNFR1, 1.9 (1.3 – 2.7) for sTNFR2, and 1.7 (1.3 – 2.3) for KIM-1. Despite the associations, each of the biomarkers alone or in combination failed to result in robust discrimination on an absolute basis or compared to a clinical model.

**Conclusion:** sTNFR1, sTNFR2, and KIM-1 were independently associated with longitudinal outcomes after discharge from a cardiac surgery hospitalization including death, cardiovascular and CKD events when obtained pre-operatively in high-risk individuals. Pre-operative plasma biomarkers could serve to assist during the evaluation of patients in whom cardiac surgery is planned.

## Introduction

Pre-existing kidney disease and post-operative acute kidney injury (AKI) are associated with poor short and long-term adverse outcomes in patients undergoing cardiac surgery.^1-3^ Although post-cardiac surgery 30-day mortality rates have decreased, annual rates remain high, exposing patients to marked sequela including cardiovascular complications and longitudinal kidney function decline.^1,3-5^ Various peri-operative risk factors have been identified in the literature.^6-9^ Nonetheless, predictive risks models have not proven to be superior to clinical judgment or have been limited by inconsistent calibration, or lack of biological data. This has limited their application among different patient cohorts and implementation in clinical practice.^4,10^ Therefore, there is an urgency to identify prognostic markers that inform on the risk for future adverse outcomes and orient on clinical strategies to prevent them.

The last decade has brought advances to non-invasive systemic and organ-specific biomarkers to assess the degree of injury and disease as well as the potential risk for complications in various clinical settings.^11^ Three of the most studied biomarkers for acute and chronic kidney disease (CKD) are the tumor necrosis factor receptors (TNFRs) and kidney injury molecule 1 (KIM-1).^12-20^ Tumor necrosis factor alpha (TNF-α) is a pleotropic cytokine that is produced predominantly by immune cells, and which can be released as a soluble circulating polypeptide.^21-23^ TNF-α binds soluble TNFR1 (sTNFR1) and sTNFR2 which in turn promote inflammatory cell recruitment, cytokine release and up-regulation of pro-fibrotic gene expression in the kidney and the heart.^24-27^ KIM-1 is expressed in the apical membrane of proximal tubular cells in response to injury and promotes kidney inflammation and fibrosis.^28,29^ Moreover, KIM-1 is expressed in injured renal tubular epithelial cells; thereby inducing the transformation of proximal tubular epithelial cells into semi-professional phagocytes. Additionally, KIM-1 has been implicated in the activation of Th1, Th2, and Th17 differentiation along with B-cell activation.^30^

Therefore, plasma assays for sTNFRs and KIM-1 may be reflective of kidney injury and immune activation, which are key pathways leading to CKD.^11^

Human studies have revealed that sTNFR1, sTNFR2, and KIM-1 are strong prognostic markers for incident and progressive CKD in persons with type 1 and type 2 diabetes mellitus in the ambulatory setting.^12,13,16,17,19,20,31,32,34,35^. Furthermore, in a previous study, our group demonstrated that 2 of these biomarkers (sTNFR1, KIM-1) were independently associated with CKD events after cardiac surgery when obtained post-operatively.^33^ However, the value of measurement of these biomarkers during the pre-operative time frame to identify patients at risk for long-term clinical sequela after cardiac surgery such as cardiovascular complications and death has not been rigorously assessed. Therefore, we sought to examine the association between pre-operative sTNFR1, sTNFR2 and KIM-1 obtained among high-risk cardiac surgery candidates and the risk for longitudinal mortality, cardiovascular events, and kidney function decline.

## Methods

### Study Design and Participants

This is an ancillary study from TRIBE-AKI, a prospective cardiac surgery cohort of 1474 adults who underwent cardiac surgery (coronary artery bypass graft [CAGB] or valve surgery). Participants undergoing elective or emergent surgery were recruited at six academic medical centers in North America between July 2007 and December 2010. Patients were at high risk for AKI, defined by the presence of one or more of the following characteristics: emergency surgery, preoperative serum creatinine > 2 mg/dl, ejection fraction < 35%, age > 70 years, diabetes mellitus, simultaneous CABG, and valve surgery, or repeat revascularization surgery. Participants with evidence of AKI before surgery, prior kidney transplantation, preoperative serum creatinine level > 4.5 mg/dl or end-stage kidney disease (ESKD) were excluded. Patients who died during the index admission (n=19) were excluded from the analysis as our main goal was to incorporate survivors and estimate the cumulative long-term risk of death as our primary endpoint. The study was approved by the institutional review board of each participating site, and written informed consent was obtained from all participants. We collected preoperative characteristics, intra-operative clinical data, and post-operative complications using the definitions of the Society of Thoracic Surgeons, and laboratory measurements in blood and urine prospectively, up to 5 years after the index admission. Full study details, including protocolized participant inclusion details and prospective clinical assessment have been described elsewhere.^36^

### Sample collection and biomarker measurement

Details of sample collection and processing have been described rigorously in previous reports.^37-40^ This study leveraged the plasma samples that were collected 2-7 days pre-operatively. Blood samples were collected in ethylenediamine tetra acetic acid tubes, centrifuged to separate plasma, and subsequently stored at -80 C. Samples underwent a single controlled thaw, were centrifuged at 5000×g for 10 minutes at 4°C, separated into 1 ml aliquots, and immediately stored at -80°C until biomarkers were measured. The plasma biomarkers sTNFR1, sTNFR2, and KIM-1 were measured using the Meso Scale Discovery platform (Meso Scale Diagnostics, Gaithersburg, MD), which uses electrochemiluminescence detection combined with patterned arrays. This technique has been implemented in several studies from TRIBE-AKI and other biomarker consortia.^13,14,16,17^ The inter-assay coefficient of variation (CV) for TNFR1 was 8.7% with a detection range of 24-100,000 pg/mL. The inter-assay CV for TNFR2 was 6.2%, with a detection range of 24-100,000 pg/mL. The inter-assay CV for KIM-1 was 16.8%, with a detection range of 42-10,582 pg/mL. We examined the association between storage time and biomarker concentration and found no significant evidence of storage effect (data not shown). Only participants with the full set of biomarkers measured pre-operatively were included in the analysis. The personnel measuring the biomarkers were blinded to clinical outcomes, including AKI, vital status, among others.

### Covariate measurement

We recorded pre-operative serum creatinine values obtained within 2 months before surgery which served to estimate baseline kidney function. The preoperative and post-operative serum creatinine measurements were performed in the same clinical laboratory for each patient at all sites. The pre-operative estimated GFR (eGFR) was obtained via the Chronic Kidney Disease Epidemiology Collaboration equation.^41^ Clinical AKI was defined as an increase of ≥ 50% or 0.3 mg/dL in serum creatinine within index hospitalization. Creatinine measurements were obtained during routine clinical care within the index hospitalization. AKI severity was ascertained via the Acute Kidney Injury Network (AKIN)^42^ staging criteria on the basis of the peak serum creatinine within the index hospitalization.

### Outcome definitions

The primary outcome for this study was long-term mortality after discharge. Vital status after the hospitalization was obtained through various mechanisms (and cross-referenced when possible). We performed phone calls to patients’ homes, searched the National Death Index, reviewed hospital records, and linkages with Center for Medicare and Medicaid Services (CMS) databases for those patients living in the United States. For Canadian participants (those enrolled into the TRIBE-AKI study in London, Ontario), we obtained data from the Registered Persons Database held at ICES to acquire vital status. These datasets were linked using unique, encoded identifiers and analyzed at the ICES. Death status and date of death were recorded accordingly.

The secondary outcomes were a) cardiovascular events and b) a composite of CKD (incidence or progression). In those individuals with an eGFR ≥ 60 mL/min per 1.73m^2^ pre-operatively, CKD incidence was defined as a 25% reduction in eGFR and a fall below 60 mL/min per 1.73m^2^. In those individuals with an eGFR < 60 mL/min per 1.73m^2^ pre-operatively, CKD progression was defined as a 50% reduction in eGFR or a fall below 15 mL/min per 1.73m^2^. These definitions were based on established cutoffs as outlined by the multi-center Assessment, Serial Evaluation, and Subsequent Sequelae in Acute Kidney Injury (ASSESS-AKI) Study.^43^ Follow-up eGFR values to ascertain the CKD outcome were only available for individuals from the Yale and London Health Science Centre sites. Cardiovascular events were defined as hospitalization for acute coronary syndrome, myocardial infarction, congestive heart failure, repeat coronary bypass, and percutaneous coronary intervention. Information about cardiovascular events was obtained via linkages with Center for Medicare and Medicaid Services (CMS) databases for patients in the United States. Variables used in the probabilistic matching to CMS data included surgery site and date, sex, race, admission date, and discharge date. Similar to the ascertainment of death among Canadian patients, clinical data was obtained from data holdings at ICES. The International Classification of Diseases (9^th^ and 10^th^ revision), Ontario Health Insurance Plan, and Canadian Classification of Health Interventions codes were used to identify patients with cardiovascular events.

### Statistical analyses

Descriptive statistics were reported as mean (SD) or median (interquartile range) for continuous variables, and as frequency (percentage) for categorical variables. When evaluating the association between biomarkers and outcomes, biomarkers were analyzed both as continuous (log_*e*_ transformed biomarker levels) and categorical (tertile) variables, with the lowest tertile serving as a reference group. Multivariable analyses were adjusted for the following variables: Age (per year), gender, white race (Yes/No), non-elective surgery, surgery type, pre-op eGFR, diabetes, hypertension, CHF, history of MI, pre-op urine albumin and urine creatinine, and clinical site. Cox proportional hazards regression models were used to examine the cause-specific association between the pre-operative biomarker measurements and the primary and secondary outcomes. Correlations between biomarkers are also presented. Kolmogorov-type supremum tests were used to evaluate proportional hazards assumptions for all models. Wilcoxon signed-rank test was used to ascertain biomarker concentrations per each outcome. In sensitivity analyses, we examined whether the presence of diabetes, as a comorbidity, was an effect modifier on the relationship between the preoperative plasma biomarkers and the outcomes. Additionally, we explored the prognostic value of these biomarkers when added to a clinical model; comprised of comorbidities and kidney-specific laboratory parameters except the 3 novel biomarkers tested. The change in the Harrell’s c-index was used to quantify the incremental value of the biomarkers considering the time-to event approach used in this study. Delong test was used to assess statistically significant differences in discrimination. Small cell counts are only presented for data collected by TRIBE-AKI and not from ICES data holdings.

For the latter, counts of 5 or fewer participants are suppressed to minimize the risk of participant re-identification. Two-tailed P values of less than 0.05 were considered statistically significant. Analyses were performed in SAS version 9.4. (SAS Institute, Cary, NC) and R 2.15.0 (R Foundation for Statistical Computing, Vienna Austria).

## RESULTS

### Baseline characteristics and post-operative AKI in TRIBE-AKI

A total of 1378 participants who underwent cardiac surgery and had the 3 pre-operative biomarkers measured were included in the analysis and CKD events were adjudicated in 837 participants (**Figure 1**). Baseline characteristics of the patients stratified by biomarker tertile (sTNFR1) are reflected in **Table 1** and **Supplemental Table 1** for sTNFR2 and KIM-1, respectively. The average age at the time of surgery was 72 ± 10 and 69% of the participants were male. Most participants were white (94%) and 83% underwent elective cardiac surgery. Those within the higher pre-operative concentrations of sTNFR1 tended to have older age, and a higher frequency of comorbidities including diabetes mellitus (44%), hypertension (84%), congestive heart failure (31%), and history of myocardial infarction (29%) as shown in **Table 1**. Similar relationships were seen for sTNFR2, and KIM-1, summarized in **Supplemental Table 1**. The mean preoperative eGFR and uACR were 68 ± 19 ml/min per 1.73 m^2^ and 0.12 ± 1.4 mg/g, respectively. Post-surgery, 34% developed AKI based upon serum creatinine metrics with the majority experiencing AKIN stage 1 (90%). Biomarker correlations included: TNFR1-TNFR2: 0.9, TNFR1-KIM1: 0.5 and KIM-1-TNFR2: 0.5.

**Figure 1.**
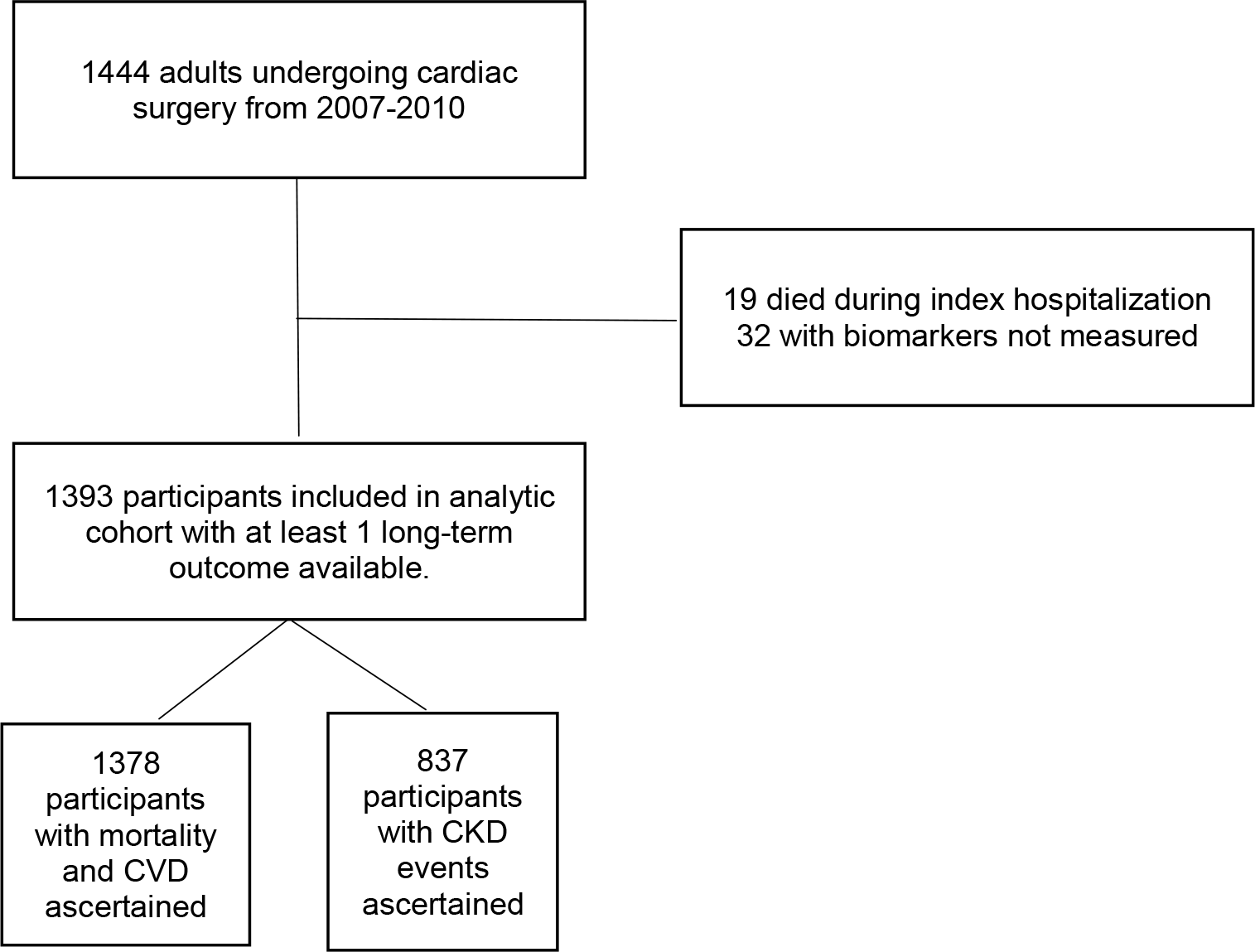
Flow chart of TRIBE-AKI analytical cohort. **Foot note:** CKD: chronic kidney disease, CVD: cardiovascular disease

**Table 1.** Pre-operative patient characteristics stratified by sTNFR1 tertiles in TRIBE-AKI. AKI, acute kidney injury defined as (0.3 mg/dl or 50% rise in serum creatinine); eGFR, estimated glomerular filtration rate calculated using CKD-EPI equation; CKD, chronic kidney disease; CABG, coronary artery bypass grafting; CPB, cardiopulmonary bypass; ICU, intensive care unit. To covert serum creatinine value from mg/dl to µmol/L please multiple by 88.4. †Albuminuria defined as Urine Albumin to Creatinine ratio >30 mg/g

|  |  | sTNFR1 |  |  |  |
| --- | --- | --- | --- | --- | --- |
| Characteristic | All (n=1393) | Pre-op Tertile 1<br>24-6467 pg/mL<br>(n=464) | Pre-op Tertile 2<br>6471-8964 pg/mL<br>(n=465) | Pre-op Tertile 3<br>8966-100000 pg/mL<br>(n=464) | P-value |
| Age, mean (at time of surgery) | 71.9 ± 9.7 | 69.2 ± 10.1 | 72.5 ± 9.8 | 73.8 ± 8.5 | <0.001 |
| White | 1,3 (94.2%) | 436 (94.0%) | 444 (95.5%) | 432 (93.1%) | 0.3 |
| Male | 963 (69.1%) | 340 (73.3%) | 321 (69.0%) | 302 (65.1%) | 0.03 |
| Diabetes | 539 (38.7%) | 165 (35.6%) | 170 (36.6%) | 204 (44.0%) | 0.02 |
| Hypertension | 1,112 (79.8%) | 348 (75.0%) | 374 (80.4%) | 390 (84.1%) | 0.003 |
| EF <35% or grade 3 or 4 LV dysfunction | 136 (9.8%) | 43 (9.3%) | 47 (10.1%) | 46 (9.9%) | 0.9 |
| Myocardial infarction | 357 (25.6%) | 104 (22.4%) | 119 (25.6%) | 134 (28.9%) | 0.3 |
| Congestive heart failure | 314 (22.5%) | 76 (16.4%) | 94 (20.2%) | 144 (31.0%) | <0.001 |
| Mean Preoperative serum Cr, mg/dL | 1.1 ± 0.3 | 0.9 ± 0.2 | 1 ± 0.21 | 1.3 ± 0.4 | <0.001 |
| Mean Preoperative eGFR,<br>mL/min/1.73m<sup>2</sup> | 68.2 ± 19.2 | 80 ± 14.4 | 70.6 ± 15.5 | 53.9 ± 17.6 | <0.001 |
| CKD stage ≥ 3 | 465 (33.4%) | 35 (7.5%) | 128 (27.5%) | 302 (65.4%) | <0.001 |
| Mean Urine Albumin to Cr ratio, mg/g | 0.1 ± 1.4 | 0.04 ± 0.10 | 0.1 ± 0.1 | 0.3 ± 2.5 | 0.03 |
| Surgery type |  |  |  |  | 0.06 |
| CABG and valve | 310 (22.3%) | 91 (19.6%) | 102 (21.9%) | 117 (25.2%) |  |
| Other | 19 (1.3%) | 8 (1.7%) | 5 (1.0%) | 6 (1.3%) |  |
| CABG only | 679 (48.7%) | 234 (50.4%) | 245 (52.7%) | 200 (43.1%) |  |
| Valve only | 385 (27.6%) | 131 (28.2%) | 113 (24.3%) | 141 (30.4%) |  |
| Nonelective surgery | 1,163 (83.5%) | 414 (89.2%) | 401 (86.2%) | 348 (75.0%) | <0.001 |

### Preoperative biomarkers and long-term mortality in TRIBE-AKI

Over a median follow-up of 6.7 (IQR 4.0-7.9) years, 434 (31%) died within the study timeframe. Pre-operatively, median levels of sTNFR1, sTNFR2 and KIM-1 were higher in participants who died during the study timeframe (**Supplementary Table 2**). The event rate for death was 53.7 per 1000 person-years (95% CI 49.0-58.8) as reflected in **Table 2** and Kaplan-Meier curves in **Figure 2**. Patients in the third tertile of sTNFR 1 (highest) exhibited approximately 2-fold the risk of all-cause mortality compared to those in the first tertile (reference group). Similarly, patients in the highest tertile of sTNFR2 and KIM-1 had 2 – 2.5-fold the risk of all-cause mortality when compared to patients in the lowest quartiles. After adjusting for clinical covariates, each natural log higher of pre-operative sTNFR1 was statistically and independently associated with long-term mortality with an adjusted HR (95% CI) of 2.9 (2.3 – 3.9), whereas each natural log higher of sTNFR2 and KIM-1 were independently associated with death with adjusted HRs (95% CI) of 2.3 (1.8 – 2.9) and 1.9 (1.6 – 2.4), respectively (**Figure 3)**. Diabetes mellitus status did not modify the association between pre-operative biomarkers and mortality (**Supplementary Table 3**).

**Table 2.** Hazard Ratios for mortality, cardiovascular and kidney events in TRIBE-AKI.

| Biomarker | Range, pg/mL<br>(Min, Max) | All-cause mortality |  | Cardiovascular Events |  | Chronic Kidney Disease Events |  |
| --- | --- | --- | --- | --- | --- | --- | --- |
|  |  | Event rate (95%CI)<br>per 1000 PY | Adjusted*<br>Hazard Ratio<br>(95%CI) | Event rate (95%CI)<br>per 1000 PY | Adjusted*<br>Hazard Ratio<br>(95%CI) | Event rate (95%CI)<br>per 1000 PY | Adjusted*<br>Hazard Ratio<br>(95%CI) |
| **sTNFR1**<br>**(pg/mL)** | Log<sub>e</sub><br>Transformed | 53.7 (49.0, 58.8) | 3.0 (2.3 – 4.0) | 34.6 (30.7, 38.9) | 2.1 (1.5 – 3.1) | 55.4 (49.1, 62.5) | 2.2 (1.5 – 3.1) |
|  | Tertile1<br>(957, 2066) | 32.2 (26.2, 39.5) | 1 (reference) | 22.9 (17.8, 29.4) | 1 (reference) | 42.0 (33.4, 52.8) | 1 (reference) |
|  | T2<br>(2067, 2829) | 44.7 (37.5, 53.2) | 1.2 (0.9 – 1.6) | 29.0 (23.2, 36.3) | 1.1 (0.8 – 1.5) | 55.5 (45.1, 68.2) | 1.4 (1.0 – 1.9) |
|  | T3<br>(2831, 4162) | 87.2 (76.3, 99.6) | 2.1 (1.5 – 2.8) | 55.0 (46.0, 65.9) | 1.7 (1.1 - 2.5) | 69.9 (56.7, 86.1) | 2.0 (1.4 – 3.0) |
| **sTNFR2**<br>**(pg/mL)** | Log<sub>e</sub><br>Transformed | 53.7 (49.0, 58.8) | 2.3 (1.8 – 2.9) | 34.6 (30.7, 38.9) | 1.9 (1.4 – 2.6) | 55.4 (49.1, 62.5) | 1.9 (1.3 – 2.7) |
|  | T1<br>(2022, 4756) | 31.4 (25.6, 38.6) | 1 (reference) | 24.7 (19.4, 31.5) | 1 (reference) | 48.4 (39.0, 60.1) | 1 (reference) |
|  | T2<br>(4758, 6651) | 47.1 (39.7, 55.8) | 1.3 (1.0 – 1.8) | 26.2 (20.6, 33.2) | 0.9 (0.6 – 1.3) | 53.8 (43.5, 66.7) | 1.1 (0.8 – 1.5) |
|  | T3<br>(6652, 9693) | 85.7 (74.9, 98.0) | 2.1 (1.6 – 2.9) | 56.0 (46.9, 66.9) | 1.6 (1.1 – 2.3) | 62.2 (50.3, 77.0) | 1.3 (0.9 – 2.0) |
| **KIM-1**<br>**(pg/mL)** | Log<sub>e</sub><br>Transformed | 53.7 (49.0, 58.8) | 2.0 (1.6 – 2.4) | 34.6 (30.7, 38.9) | 1.6 (1.2 – 2.1) | 55.4 (49.1, 62.5) | 1.7 (1.3 – 2.3) |
|  | T1<br>(42.0, 205.3) | 24.7 (19.6, 31.1) | 1 (reference) | 18.0 (13.6, 23.7) | 1 (reference) | 36.8 (28.8, 47.1) | 1 (reference) |
|  | T2<br>(205.4, 290.0) | 54.9 (46.8, 64.3) | 1.8 (1.3 – 2.4) | 39.3 (32.2, 47.9) | 1.9 (1.3 – 2.7) | 55.7 (45.2, 68.7) | 1.6 (1.1 – 2.2) |
|  | T3<br>(290.2, 8599.8) | 85.3 (74.5, 97.7) | 2.7 (2.0 – 3.7) | 49.8 (41.2, 60.1) | 2.1 (1.4 – 3.0) | 75.0 (61.7, 91.1) | 2.2 (1.6 – 3.2) |

\*Adjusted for recipient Age (per year), recipient gender, white race (Yes/No), non-elective surgery, surgery type, pre-op eGFR, diabetes, hypertension, CHF, history of MI, pre-op urine albumin and urine creatinine, clinical site

**Figure 2.**
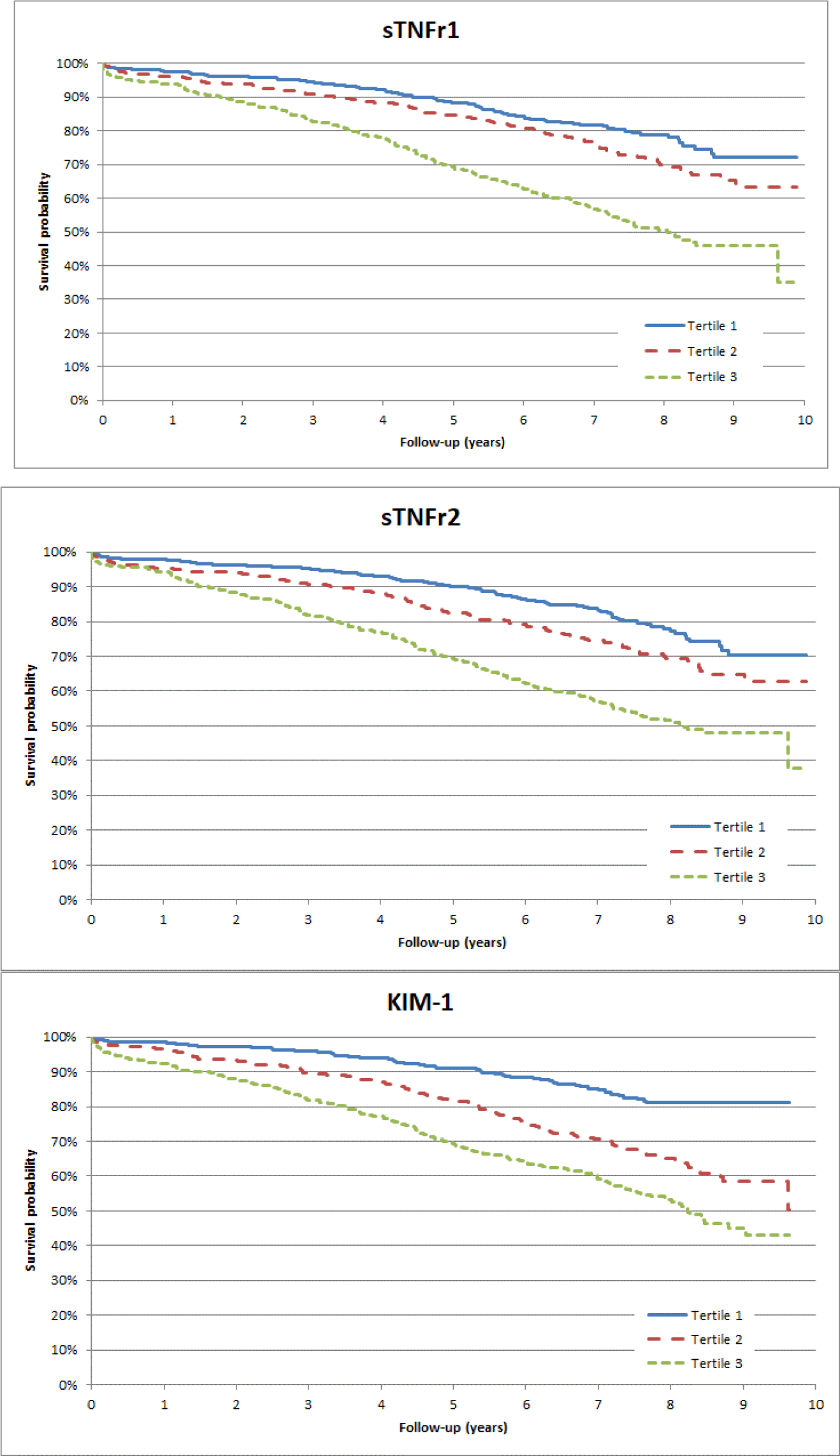
Kaplan-Meier curves for mortality for each biomarker tertile.

**Figure 3.**
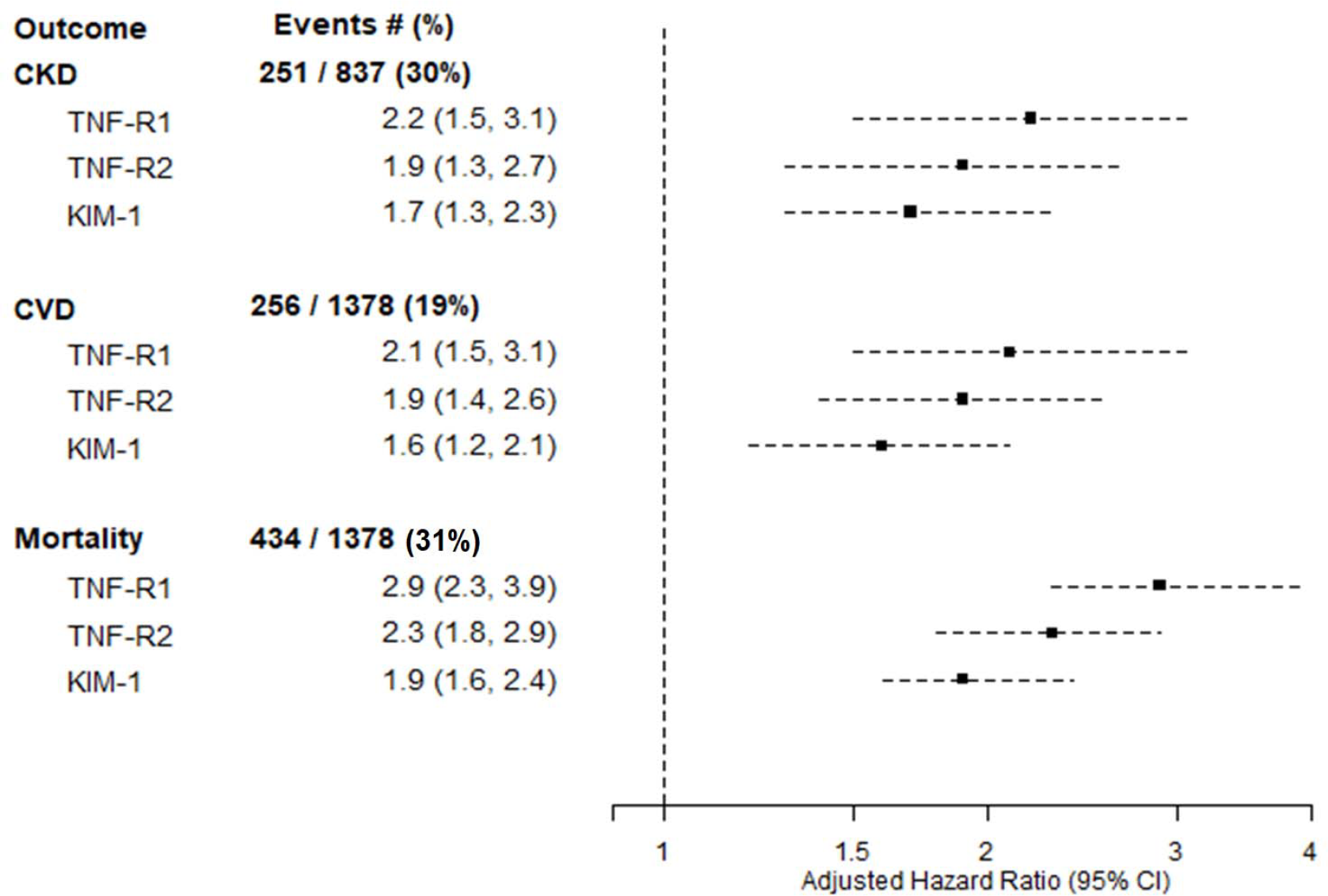
Plasma biomarkers and hazard ratios for all-cause mortality, chronic kidney disease events, and cardiovascular events. Hazard ratios were adjusted for age (per year), sex, white race (yes/No), non-elective surgery, surgery type (CABG, valve, combined), pre-op eGFR, history of diabetes, history of hypertension, CHF, history of MI, pre-op urine albumin and urine creatinine, and center.

### Preoperative biomarkers and cardiovascular events in TRIBE-AKI

Over a median follow-up of 6.3 (IQR 3.1-7.7) years, 256 (19%) participants had a cardiovascular event. At baseline, median levels of sTNFR1, sTNFR2 and KIM-1 were higher in participants who had cardiovascular events compared to those who did not within the study timeframe (**Supplementary Table 2**). Patients in the highest tertile of sTNFR1 had 1.7-fold risk (95% CI: 1.1-2.5) of cardiovascular events compared to those in the lowest tertile. Similarly, patients in the highest tertiles of sTNFR2 and KIM-1 had 1.6 – 2-fold risk of cardiovascular events compared to those in the lowest tertile as shown in **Table 2**. After adjusting for key covariates, each natural log higher of sTNFR1, sTNFR2 and KIM-1 was statistically and independently associated to cardiovascular events with aHRs (95% CI) of 2.9 (2.3 – 3.9), 2.3 (1.8 – 2.9) and 1.9 (1.6 – 2.4), respectively (**Figure 3**). There was no interaction by diabetes mellitus status for the association between any of the biomarkers and cardiovascular events (**Supplementary Table 3**).

### Preoperative biomarkers and long-term kidney events in TRIBE-AKI

During a median follow-up of 5.7 years (IQR: 4.2 – 7.1), 251 (30%) of 837 participants with follow-up kidney function measurements developed incident or progressive CKD. Characteristics between those that had data on long-term kidney function vs. those without were largely similar (**Supplementary table 4**). Baseline median levels of sTNFR1, sTNFR2 and KIM-1 were higher in participants who had kidney events compared to those who did not (**Supplementary Table 2**). Patients in the highest tertile of sTNFR1, sTNFR2 and KIM-1 had between 1.7-2.2-fold risk of kidney events compared to patients in the lowest tertile (**Table 2**). In adjusted models, each natural log higher of baseline concentration of sTNFR1, sTNFR2 and KIM-1 was associated to kidney events with aHRs (95% CI) of 2.2 (1.5 – 3.1), 1.9 (1.3 – 2.7) and 1.7 (1.3 – 2.3). There was no interaction by diabetes mellitus status with the association between biomarkers and kidney events (**Supplementary Table 3**). Addition of the biomarkers to the full clinical model, individually or all 3 combined, resulted in negligible differences in discrimination for each of the outcomes as measured by c-statistics that were all generally < 0.70 (**Table 3**).

**Table 3.** C-index of Clinical Variables and Biomarkers.

| Outcome | Model |  |  |  |  |
| --- | --- | --- | --- | --- | --- |
|  | Clinical Alone | Clinical + sTNFR1 | Clinical + sTNFR2 | Clinical + KIM1 | Clinical + All 3 Markers |
| CKD events | 0.626 | 0.648 | 0.635 | 0.644 | 0.652 |
| CVD | 0.669 | 0.678 | 0.676 | 0.689 | 0.692 |
| Death | 0.667 | 0.685 | 0.689 | 0.696 | 0.703 |

CKD: chronic kidney disease; CI: confidence interval; CVD: cardiovascular disease
Clinical model= Age (per year), recipient gender, white race (Yes/No), non-elective surgery, surgery type, pre-op eGFR, diabetes, hypertension, CHF, history of MI, pre-op urine albumin and urine creatinine, and center

## DISCUSSION

In this study, we examined the association of biomarkers of kidney inflammation and injury obtained prior to cardiac surgery to inform on the risk of long-term clinical events among patients at high-risk for AKI. We demonstrated that high pre-operative levels of sTNFR1, sTNFR2, and KIM-1 were strongly associated with mortality, cardiovascular, and kidney events after adjusting for key clinical covariates within 6 years of follow-up. Furthermore, diabetes mellitus status did not modify the association between pre-operative biomarkers and the aforementioned outcomes, which is a novel observation in the field of biomarkers and CKD. Our findings are concordant with those obtained in other clinical inpatient and outpatient settings and support the key role of these pathways in the development of kidney and cardiovascular sequela in multiple heterogeneous populations.^12,16,17,20,37,44-50^

The novel aspect of our study is that we showed the association of these biomarkers before a major clinical stressor was applied, highlighting their potential use during the pre-cardiac surgery period. Patients in the highest tertile of sTNFR1, sTNFR2 and KIM-1 experienced 2 to 2.7-fold increase in the risk of all-cause of mortality when this outcome as ascertained within 6 years. Signals of the same magnitude were seen on their association with longitudinal cardiovascular events. There are several plausible mechanisms that could explain these findings. TNF – α signaling pathways are key mediators of the inflammatory milieu in the heart, kidney, and other organs.^24,25,28,29,51^ In cardiomyocytes, activation of TNFR1 and TNFR2 evokes different and complementary effects. While activation of TNFR1 results in the generation of reactive oxygen species (ROS), inflammatory cell recruitment and upregulation of cytokines, TNFR2 promotes downstream effects including phosphorylation of the extracellular signal-regulated kinase (ERK), protein kinase C delta and calmodulin kinase which are involved in ischemia/reperfusion injury and protection from it.^52^ Additional knockout mouse models revealed that silencing TNFR1 expression optimizes cardiac performance and reduces infarct size during ischemia/reperfusion injury while left ventricular dysfunction and dilatation are exaggerated when repressing TNFR2 genes.^52-55^ Soluble TNFRs, more stable markers of chronic inflammation, have been studied among patients with stable coronary artery disease. In accordance with our findings, sTNFRs have been strongly and independently associated with heart failure and cardiovascular events in patients with and without kidney disease.^45,56-58^ However, their ability to associate with such adverse outcomes many years after cardiac surgery has not been reported before and these findings require further investigation.

In the kidney, sTNFR1 and sTNFR2 are expressed in the glomeruli, endothelial and tubular cells, and both coordinate downstream cellular responses that lead to apoptosis, and generation of proinflammatory mediators that are responsible of severe kidney injury.^11,25,26^ Data supporting the association of these biomarkers with progression of kidney disease among stable CKD patients with and without diabetes mellitus have been reported elsewhere.^12,19,32,50,58,59^ In this study, each natural log higher concentration of sTNFRs before surgery was associated with nearly 2-fold increase in the risk of long-term kidney events. KIM-1 (in urine and plasma) has been widely recognized as an early biomarker of AKI and CKD in models of drug-induced nephrotoxicity, ischemia-reperfusion, among others.^28,29,60-62^ While kidney tubules have remarkable capacity to self-repair following injury, pathological processes can predominate and therefore, incremental levels of KIM-1 could serve as a marker of subclinical or ongoing tubular injury which is defined biological pathway leading to CKD/ESKD.^11,28,29,39,62^ Therefore, as demonstrated in clinical studies, patients with higher levels of KIM-1 (compared to those with lower levels) experience longitudinal function decline and disease progression as demonstrated in CKD patients with and without diabetes mellitus.^13,17,20,35,48,49^ In contrast to sTNFRs, KIM-1 is primarily expressed in the proximal tubules and has been markedly associated with kidney-related outcomes and mortality, but not cardiovascular disease. There is a paucity of studies exploring the role of plasma KIM-1 in cardiovascular outcomes among kidney disease patients.

Urinary KIM-1 levels have not demonstrated to be associated to longitudinal cardiac events in the Health, Aging and Body Composition (Health ABC) study.^47^ Similarly, our group has previously demonstrated through mediation analyses that elevated urinary KIM-1 levels, while associated with all-cause mortality and CKD outcomes after post-operative AKI, were not associated with cardiovascular disease after cardiac surgery.^46^ It is accepted that cardiovascular disease is a competing risk in AKI/CKD and share common pathophysiological pathways such as TNF – α/TNFR^63^ and therefore, underlying inflammation and kidney tubule injury may represent a separate pathway.^33^

Our study has several strengths, including our large sample size which allowed us to control for important covariates that could compromise the association of biomarkers with outcomes. To the best of our knowledge, this is the first study that ascertain the association of well-characterized biomarkers (sTNFR1, sTNFR2 and KIM-1) obtained prior to a clinical insult and long-term post-operative kidney and non-kidney related outcomes. Additionally, we ascertained mortality status in 99% of study participants. In terms of limitations, we were only able to ascertain CKD outcomes post-surgery in two of the participating institutions, and the lack of kidney events could have compromised our analyses. Furthermore, this study did not account for post-index surgery hospitalizations or visits to the emergency department after discharge which could arguably modify the risk of subsequent clinical events. It is also possible that capturing only 2 pathways of kidney disease (injury and inflammation) remains a limited approach to predict kidney disease progression. Additional biomarkers may be needed to improve discrimination for long-term outcomes post-surgery. Lastly, our study participants were predominantly white, and this study lacks a validation cohort, both of which could limit the generalizability of our results.

In conclusion, pre-operative concentrations of plasma sTNFR1, sTNFR2, and KIM-1 were associated with long-term cardiovascular complications, kidney events, and death after cardiac surgery in a high-risk population for kidney disease and complications. Diabetes mellitus did not modify the association between these biomarkers and all three clinical outcomes. Future studies should examine the utility of measuring plasma sTNFR1, sTNFR2, and KIM-1 in other surgical settings.

## Supporting information

Strobe check list for cohort studies

## Data Availability

All data produced in the present study are available upon reasonable request to the authors

## Authors’ contributions

GVR and SGC drafted the manuscript. HTP and EMA provided expert statistical consultation. GVR, HTP, EMA, and CRP analyzed the data. All authors provided meaningful intellectual contributions throughout the elaboration of the manuscript and accepted the final version of it.

## Acknowledgements

The research reported in this article was supported by the American Heart Association Clinical Development Award, as well as by the grant R01HL-085757 from the National Heart, Lung, and Blood Institute. C.R.P. is also supported by an NIH grant (K24DK090203). S.G.C., A.X.G., and C.R.P. are also members of the NIH-sponsored Assess, Serial Evaluation, and Subsequent Sequelae in Acute Kidney Injury (ASSESS-AKI) Consortium (U01DK082185). S.G.C. M.G.S, and C.R.P. are members and are supported in part by the Chronic Kidney Disease Biomarker Consortium (1U01DK106962-01). DGM is supported by K23DK117065. JLK is supported by R21DK113420. The study was also supported by CTSA Grant Number UL1 RR024139 from the National Center for Research Resources (NCRR). This study was supported by the Institute for Clinical Evaluative Sciences (ICES), which is funded by an annual grant from the Ontario Ministry of Health and Long-Term Care (MOHLTC); Dr. Amit Garg is supported by the Dr. Adam Linton Chair in Kidney Health Analytics; no other relationships or activities that could appear to have influenced the submitted work. S.G.C. and C.R.P. are on the Advisory Board of RenalytixAI, and both own equity in the same. S.G.C. has served as a consultant for AKI therapeutics for Quark Biopharma and CHF Solutions. The opinions, results and conclusions reported in this paper are those of the authors and are independent from the funding sources. No endorsement by ICES or the Ontario MOHLTC is intended or should be inferred. Parts of this material are based on data and information compiled and provided by CIHI. However, the analyses, conclusions, opinions, and statements expressed herein are those of the authors, and not necessarily those of CIHI. All other authors have reported that they have no relationships relevant to the contents of this paper to disclose.

---

**Supplemental Table 1.** Patient characteristics stratified by sTNFR2 and KIM-1 tertile in TRIBE-AKI

| Characteristic | sTNFR2 |  |  |  | KIM-1 |  |  |  |
| --- | --- | --- | --- | --- | --- | --- | --- | --- |
|  | Pre-op Tertile 1<br>(24-4284)<br>(n=464) | Pre-op Tertile 2<br>(4290-5918)<br>(n=465) | Pre-op Tertile 3<br>(5935-100000)<br>(n=464) | P-value | Pre-op Tertile 1<br>(42-205)<br>(n=464) | Pre-op Tertile 2<br>(205-290)<br>(n=465) | Pre-op Tertile 3<br>(290-8600)<br>(n=464) | P-value |
| Age, mean (at time of surgery) | 69.5 ± 10.2 | 72.3 ± 9.7 | 73.7 ± 8.8 | <0.001 | 69.5 ± 10.8 | 73.5 ± 8.7 | 72.5 ± 9.1 | <0.001 |
| White Race | 436 (94.0%) | 444 (95.5%) | 432 (93.1%) | 0.29 | 437 (94.2%) | 445 (95.7%) | 430 (92.7%) | 0.1 |
| Male sex | 349 (75.2%) | 310 (66.7%) | 304 (65.5%) | 0.002 | 325 (70.0%) | 321 (69.0%) | 317 (68.3%) | 0.9 |
| Diabetes | 166 (35.8%) | 174 (37.4%) | 199 (42.9%) | 0.07 | 144 (31.0%) | 160 (34.4%) | 235 (50.6%) | <0.001 |
| Hypertension | 348 (75.0%) | 372 (80.0%) | 392 (84.5%) | 0.002 | 347 (74.8%) | 377 (81.1%) | 388 (83.6%) | 0.003 |
| EF <35% or grade 3 or 4 LV dysfunction | 43 (9.3%) | 45 (9.7%) | 48 (10.3%) | 0.86 | 42 (9.1%) | 45 (9.7%) | 49 (10.6%) | 0.7 |
| Myocardial infarction | 102 (22.0%) | 117 (25.2%) | 138 (29.7%) | 0.044 | 116 (25.0%) | 109 (23.4%) | 132 (28.4%) | 0.4 |
| Congestive heart failure | 77 (16.6%) | 95 (20.4%) | 142 (30.6%) | <0.001 | 85 (18.3%) | 99 (21.3%) | 130 (28.0%) | 0.001 |
| Mean Preoperative serum Cr, mg/dL | 0.9 ± 0.2 | 1.0 ± 0.2 | 1.3 ± 0.4 | <0.001 | 0.9 ± 0.2 | 1.0 ± 0.3 | 1.2 ± 0.4 | <0.001 |
| Preoperative eGFR, mL/min/1.73m<sup>2</sup> (mean ± SD) | 79.6 ± 14.6 | 70.2 ± 15.7 | 54.8 ± 18.3 | <0.001 | 75.0 ± 16.3 | 68.9 ± 17.7 | 60.6 ± 20.6 | <0.001 |
| CKD stage ≥ 3 | 41 (8.8%) | 132 (28.4%) | 292 (62.9%) | <0.001 | 87 (18.7%) | 139 (29.9%) | 239 (51.5%) | <0.001 |
| Urine Albumin to Cr ratio, mg/g (mean±SD) | 0.04 ± 0.1 | 0.1 ± 0.1 | 0.3 ± 2.5 | 0.04 | 0.03 ± 0.1 | 0.2 ± 0.1 | 0.3 ± 2.5 | 0.03 |
| Surgery type |  |  |  | 0.13 |  |  |  | 0.02 |
| CABG and valve | 85 (18.3%) | 108 (23.2%) | 117 (25.2%) |  | 90 (19.4%) | 99 (21.3%) | 121 (26.1%) |  |
| Other | 9 (1.9%) | 4 (0.9%) | 6 (1.3%) |  | 2 (0.4%) | 12 (2.6%) | 5 (1.1%) |  |
| CABG only | 242 (52.2%) | 230 (49.5%) | 207 (44.6%) |  | 247 (53.2%) | 216 (46.5%) | 216 (46.6%) |  |
| Valve only | 128 (27.6%) | 123 (26.5%) | 134 (28.9%) |  | 125 (26.9%) | 138 (29.7%) | 122 (26.3%) |  |
| Nonelective surgery | 405 (87.3%) | 402 (86.5%) | 356 (76.7%) | <0.001 | 389 (83.8%) | 401 (86.2%) | 373 (80.4%) | 0.05 |

**Supplementary Table 2.** Biomarker Concentrations by each outcome.

|  | All-cause mortality |  |  | Cardiovascular event |  |  | Chronic Kidney Disease events |  |  |
| --- | --- | --- | --- | --- | --- | --- | --- | --- | --- |
|  | Died | Survived | P-value | Yes | No | P-value | Yes | No | P-value |
| **sTNFR1**<br>**(pg/mL)** | 8937<br>(6696-12708) | 7038<br>(5740-9088) | <0.001 | 7540<br>(6278-9965) | 7036<br>(5710-9082) | <0.001 | 8566<br>(6508-12226) | 7303<br>(5939-9774) | 0.002 |
| **sTNFR2**<br>**(pg/mL)** | 5817<br>(4539-8418) | 4765<br>(3764-6044) | <0.001 | 5040<br>(3957-6728) | 4889<br>(3792-6343) | <0.001 | 5711<br>(4188-8113) | 4927<br>(3873-6456) | 0.06 |
| **KIM-1**<br>**(pg/mL)** | 284.1<br>(228.2-379.6) | 223.8<br>(173.1-296.5) | <0.001 | 269.6<br>(204.9-335.2) | 227.0<br>(176.6-299.4) | <0.001 | 276.3<br>(215.2-371.5) | 239.4<br>(180.9-316.3) | <0.001 |

Median (IQR) values are presented in each cell.

**Supplementary Table 3.** Assessment of the interaction by diabetes mellitus on the association between pre-operative biomarkers and long-term outcomes in TRIBE-AKI

| Biomarker (log) | Diabetes Mellitus status | All-Cause Mortality |  | Cardiovascular Events |  | Chronic Kidney Disease Events |  |
| --- | --- | --- | --- | --- | --- | --- | --- |
|  |  | Adjusted* HR (95% CI) | Interaction p-value | Adjusted* HR (95% CI) | Interaction p-value | Adjusted* HR (95% CI) | Interaction p-value |
| **sTNFR1** | No | 2.3 (1.8, 3.7) | 0.5 | 1.9 (1.1, 3.1) | 0.9 | 1.9 (1.1, 3.2) | 0.2 |
|  | Yes | 3.6 (2.4, 5.5) |  | 2.3 (1.3, 3.9) |  | 2.9 (1.8, 4.7) |  |
| **sTNFR2** | No | 2.1 (1.6, 2.8) | 0.8 | 1.9 (1.3, 2.8) | 0.5 | 1.5 (0.9, 2.5) | 0.09 |
|  | Yes | 3.5 (2.2, 5.5) |  | 1.8 (1.0, 3.2) |  | 2.8 (1.6, 4.9) |  |
| **KIM-1** | No | 2.6 (1.8, 3.7) | 0.07 | 1.6 (0.9, 2.5) | 0.9 | 2.2 (1.3, 3.8) | 0.8 |
|  | Yes | 1.8 (1.4, 2.5) |  | 1.7 (1.2, 2.4) |  | 1.7 (1.2, 2.5) |  |

\*Adjusted for recipient Age (per year), recipient gender, white race (Yes/No), non-elective surgery, surgery type, pre-op eGFR, diabetes, hypertension, CHF, history of MI, pre-op urine albumin and urine creatinine, clinical site

**Supplementary Table 4.** Pre-operative patient characteristics stratified by availability of CKD outcome assessment in follow-up

| Characteristic | CKD outcome available |  |
| --- | --- | --- |
|  | No (n=556) | Yes (n=837) |
| Age, mean (at time of surgery) | 71.9 ± 10.8 | 71.8 ± 8.9 |
| White | 503 (90.5%) | 809 (96.7%) |
| Male | 370 (66.5%) | 593 (70.8%) |
| Diabetes | 202 (36.3%) | 337 (40.3%) |
| Hypertension | 452 (81.3%) | 660 (78.9%) |
| EF <35% or grade 3 or 4 LV dysfunction | 71 (12.8%) | 65 (7.8%) |
| Myocardial infarction | 146 (26.3%) | 211 (25.2%) |
| Congestive heart failure | 209 (37.6%) | 105 (12.5%) |
| Mean Preoperative serum Cr, mg/dL | 1.1 ± 0.4 | 1.0 ± 0.3 |
| Mean Preoperative eGFR, mL/min/1.73m<sup>2</sup> | 65.8 ± 20.3 | 69.8 ± 18.3 |
| CKD stage ≥ 3 | 215 (38.7%) | 250 (29.9%) |
| Mean Urine Albumin to Cr ratio, mg/g | 0.2 ± 2.2 | 0.1 ± 0.2 |
| Surgery type |  |  |
| CABG and valve | 125 (22.5%) | 185 (22.1%) |
| Other | 10 (1.8%) | 9 (1.0%) |
| CABG only | 263 (47.3%) | 416 (49.7%) |
| Valve only | 158 (28.4%) | 227 (27.1%) |
| Nonelective surgery | 400 (71.9%) | 763 (91.2%) |

